# Association of maternal hepatitis B virus infection with adverse pregnancy outcomes and prenatal screening result of second-trimester: a retrospective cohort study

**DOI:** 10.1101/2020.04.20.20068874

**Authors:** Quanze He, You Zhou, Xiaojuan Wu, Ying Xue, Chunhua Zhang, Lu Lu, Hankui Liu, Jianguo Zhang, Xiao Dang, Ting Wang, Hong Li

## Abstract

**Objective:** The association between maternal HBV infection and adverse pregnancy outcomes remains controversial and the prenatal screening features was not investigated though they strongly indicate adverse pregnancy outcomes. We conducted a retrospective cohort study to determine the association between maternal HBV infection and adverse pregnancy outcomes and the result of prenatal screening.

**Design:** The information from 65,257 pregnant women who performed non-invasive prenatal screening (NIPS) in second-trimester from July 2015 to Nov 2019 in Suzhou, China were collected. The participants were divided into the group of “control” (n = 63,591) and “exposure” (n = 1,666). Meanwhile, eight types of adverse pregnancy outcomes in history and twelve prenatal screening results in the second-trimester of current pregnancy were investigated by estimating and adjusting their risk ratios (aRR) for women between HBV infected and uninfected using multivariate logic regression.

**Result:** Our results suggested that women infected with HBV have higher risks on the biochemical pregnancy, extrauterine pregnancy and three kinds of screen result for fetal Down’s syndrome (DS) than women with HBV un-infected. Meanwhile, the diagnostic result for the positive result of fetal Down syndrome in NIPS suggested the 98% additional risk in women with HBV infected than uninfected and supported by all adjusted models.

**Conclusion:** Taken together, maternal HBV infection is an independent risk factor of biochemical pregnancy, extrauterine pregnancy, and fetal Down’s syndrome. It also indicates the influence of maternal HBV infection way across all gestation.

## Introduction

HBV infection is a common health problem in the world, causing more than 250 million individuals became chronic carriers and 880 thousand people died from serious hepatocellular diseases with HBV chronic infection such as cirrhosis and hepatocellular carcinoma^1^. In China, the total carrier rate of HBV is around 8%, and the carrier rate of women during reproductive age and pregnancy is between 3.87–9.98%, which are higher than other countries in the world^2,3^. More importantly, the genetic material of HBV could be naturally integrated into host genetic^3-5^ and its antibody or DNA detected in ovarian tissues, placental, and ovum^6-8^. These results indicated that the influence of maternal HBV infection may across all the stages of pregnancy and result in unpredicted pregnancy outcomes, especially in the early development of fetus. Additionally, contact with blood or other body fluids in childbirth is also an important transmission channel of HBV from mother to child^1,9-11^. Thus, to clarify the potential pregnant risks of women with HBV infection is urgent for reduction the pregnant damage and protection the health of woman and child.

In previous studies, maternal HBV infection has been considered a risk factor of adverse pregnancy outcomes such as low birth weight, miscarriage, preterm birth, pre-eclampsia, gestational diabetes mellitus, postpartum hemorrhage, intrahepatic cholestasis, cesarean section and placenta previa^12-17^, but some studies reported inconsistent results^2,18-21^, even in the cohort studies with large sample size^13,18^. Furthermore, previous studies mainly focused on discussing the association between maternal HBV infection and adverse pregnancy outcomes but rarely investigated the association between maternal HBV infection and the prenatal screening results even their results indicate the potential risk of adverse pregnancy outcomes, such as ultrasound and Down syndrome screening. Actually, the abnormal results of prenatal screening were indications the potential risk of adverse pregnancy outcomes. In the first-trimester, the low progesterone, loss fetal heart sound and abnormal size of pregnancy bursa were alarms of early spontaneous abortion. In the second-trimester, the most of structural abnormal of fetus were direct reasons of miscarries such as tetralogy of fallot, malformation of appendicular skeleton and cleft lip and palate. The soft markers of ultrasound in second-trimester also suggested the potential risk of miscarries such as hypoplastic nasal bone or increased nuchal translucency (NT) or nuchal fold (NF) (I-NTF). The two soft markers of ultrasound were indicated the fetal risk on Down syndrome (DS) and their detection ratio were over 40% ^22,23^. Multiples serum markers (MSM) also used to estimate the fetal risk of Down syndrome(DS), Edward syndrome (ES) and neural tube defect (NTD) by proteins quantification of ß-HCG, alpha-fetoprotein (AFP) in maternal serum and combined several factors (such as age, gestation weeks and the information of DS and ES event in history) ^24,25^. Mild ventriculomegaly in prenatal is a mark to predict the abnormal development of the neonatal brain^26^. They were the reasons of miscarriage (spontaneous abortion or abortion) in second-trimester. Thus, using prenatal screening results to investigate the pregnant risk of women with HBV infection is feasible, and they not only provided measurable scale to indicate potential adverse pregnancy outcomes in biochemistry or image, but also demonstrate an elaborate landscape of adverse pregnancy outcomes.

In the second-trimester, the screening result of multiples serum markers, ultrasound and NIPS were importantly clinical marks for estimation fetal health and were routinized prenatal screening. NIPS is the newest method to estimate the DS, ES and Patau syndrome (PS) risk of fetus by sequencing the cell-free fetal DNA (cffDNA) in maternal plasma and has higher accuracy than MSM and ultrasound. It has been widely accepted by pregnant women in clinical though the detected types of fetal abnormal is smaller than ultrasound. Ultrasound is the most commons screening method in the three kinds of screening method and be used in detection multiple tissue deformity of fetus such as heart, skeleton, brain, live, kidney, vascellum, nuchal translucency (NT) and nuchal fold (NF).

In this study, the information from 65,257 pregnancy women were collected to investigate the association between maternal HBV infection with eight kinds of adverse pregnancy outcomes and 12 kinds of prenatal screening results in second-trimester, who performed NIPS from July 2015 and Nov 2019 were collected and 1,666 pregnancy women with HBV infected contained. The eight kinds of adverse pregnancy outcomes were classified according to the prenatal history of participant and the prenatal screening results were from their current pregnancy. Our result suggested that the fetal had a higher risk of Down syndrome in the women with HBV infection and the risk of biochemical pregnancy and extrauterine pregnancy also significantly increased in the women of HBV infected than un-infected.

## Methods

### Participants

This project was approved by the Reproductive Medicine Ethics Committee of Suzhou Municipal Hospital and launched in July 2015(approval number K901001). According to the NIPS guideline of ACMG, pregnant women were approved except whose gestation week was under 12 week, or these situations occurred: structural abnormalities of fetus, chromosomal abnormal in spouse, family history of genetic diseases, cancer, organ transplantation, stem cell treatment, blood transfusion (in last four weeks), and so on^27^. The documents of NIPS application were submitted after in-depth communication and assessment by the genetic doctor and stored in a web system. The adverse pregnancy outcomes in history and prenatal screening results in the current pregnancy were included in the application documents. All participants were followed up by hospital staff after the next ten months. In our cohort, 63,591 out 65,257 (97.5%) pregnant women were not infected by HBV (HBV-N), only 2.5% (1,666/65,257) were infected (HBV-P). Totally, 299 (0.46%) were reported as high risk of DS in NIPS in which 241 (0.37%) confirmed by amniocentesis.

### Procedures

According to the definite clinical diagnostic and prenatal screening results of pregnant women, six kinds of population characteristics, eight kinds of adverse pregnancy outcomes and 12 kinds of prenatal screening results were extracted. Population characteristics include the maternal age (year), maternal weight (kg), maternal height (cm), body mass index, (kg/m2), the count of childbirth. Meanwhile, the proportion of pregnant women in three different stages of age were also calculated (lower 30 years, between 30 and 35 years and over 35 years). Eight kinds of adverse pregnancy outcomes were classified into three classes: miscarriage, abnormal pregnancy, and other adverse pregnancy outcomes. In the class of miscarriage, four common events of fetal loss were included: development stopped on embryonic stage (before the 9th week after conception or week 11 after the last menstrual period), development stopped on fetus (over 9th week after conception), abortion and spontaneous abortion. Molar pregnancy, biochemical pregnancy, and extrauterine pregnancy constructed the class of abnormal pregnancy. The pregnant women, who lack the detailed information about the cause of adverse pregnancy outcomes, were included in the class of other adverse pregnancy outcomes.

The twelve prenatal screening results from three prenatal screening testing in the second-trimester were used and their detection methods were different: MSM (protein quantification), NIPS (genetic sequencing) and ultrasound (image). In MSM, the quantification of AFP in maternal serum used to estimate the fetal risk of DS, if the risk value over the local cut off risk value (1/300) then reported the high risk of fetal DS. Similarly, the ßHCG was used to estimate the fetal risk of ES if the risk value over 1/375 (local cut off risk value) then reported the high risk of fetal ES. The high risk of fetal NTD was reported if the quantity level of AFP over 2.5 times multiples of the median (MoM) of AFP. Here, the high risk of DS, ES and NTD named by HRDS-T, HRES-T and HRNTD, respectively.

Ultrasound as routine screening used to detect various fetal anomaly across the gestation. According to fetal ultrasound result in the current pregnancy, six kind of ultrasound abnormal were extracted: fetal multiple malformations, choroid plexus cysts, echogenic intracardiac focus, I-NTF (increased nuchal translucency (NT) or nuchal fold (NF)), SADKI (single umbilical artery or abnormal development on kidney, intestine) and others. Previous studies suggested that I-NTF is associated with DS^25,28^ and if the NT thickness of fetus is over 2.5 millimeters (mm) (gestation weeks between 10 and 14), over the 95% quantile ^29^ or the NF thickness of fetus over 5mm in the gestation weeks between 16 to 18 or over 6mm in the gestation week between 19 to 24^30,31^.

NIPS is the most popular method of screening fetal DS, ES, and PS by sequencing the cffDNA (cell free fetal DNA) from maternal plasma. The high risk of DS, ES and PS were identified if the Z score of chromosomes 21, 18 and 13 respectively over 3^32^ and their results assessed as potential adverse pregnancy outcomes. Here, they were named HRDS-N, HRES-N and HRPS-N, respectively. Finally, the variables of maternal age, maternal weight, maternal high, BMI and the count of childbirth were continuous variables, while others were logic variables and marked by “Yes” or “No” for each participant.

### Statistical analysis

In the groups of HBV-P and HBV-N, their distribution characteristic of population, adverse pregnancy outcomes, prenatal screening result and summaries were demonstrated using mean, minima, maxima, number and proportion, respectively. The characteristic of continuous variables distribution shown by mean, minima and maxima and shown the styles in Table 1 as following: mean (minima-maxima). The distribution different of maternal age, maternal weight, maternal height, BMI, and the count of pregnancy and childbirth between HBV-P and HBV-N estimated by T-test. The characteristic of logic variables shown by number and proportion and set the style in Table 1 as following: number (proportion). Their distribution different estimated by χ2 test or Fisher’s exact test. If any number in 2 × 2 chi-squared is lower 4 then Fisher’s exact test extracted (HRES-N, HRPS-N, and molar pregnancy) otherwise χ2 test performed (Table 1). The risk ratio (RRs) and confidence intervals at 95% (95%CIs) of women with HBV infection than un-infection on adverse pregnancy outcomes and prenatal screening results were assessed and adjusted (aRRs) by three multivariate logistic regression models. In model A, the covariates were the age, weight, height, BMI of the pregnant women and the number of childbirths. In model B, the covariates in model A were used but the pregnant women age replaced by the stages of age (lower 30 years, between 30 and 35 years, over 35 years) and marked “1”, “2” and “3”, respectively. In model C, the covariates in model B were used but the number of childbirths replaced by “childbirth” and “un-childbirth”. Finally, the full model performed to adjust the RRs of the 12 prenatal screening results in current pregnancy using the covariates of model C and all investigated adverse pregnancy outcomes. The analysis tool is R package (version 3.6) and the p values less than 0.05 with two-sided was deemed to be statistically significant.

**Table 1:** Classification and proportion of population characteristics, adverse pregnancy outcomes in history and prenatal screening results in current pregnancy in HBV-P and HBV-N group.

| Population characteristics |  | All (65,257) | HBV un-infected group (HBV-N) (n=63,591) | HBV infected group (HBV-P) (n=1,666) | p value |
| --- | --- | --- | --- | --- | --- |
| Maternal age, (year) <sup>1</sup> |  | 30.4 (15-54) | 30.4 (15-54) | 32 (15-45) | 0.28 <sup>3</sup> |
| 15 - 29 <sup>2</sup> |  | 30,104 (46.13%) | 29,568 (46.50%) | 536 (32.17%) | 1.1E-42 <sup>4</sup> |
| 30 - 35 <sup>2</sup> |  | 23,929 (36.79%) | 23,266 (36.59%) | 663 (39.8%) |  |
| > 35 <sup>2</sup> |  | 11,224 (17.20%) | 10,757 (16.92%) | 467 (28.03) |  |
| Maternal weight, (kg) <sup>1</sup> |  | 58.4 (30-166) | 58.3 (30-166) | 58.9 (37-132) | 1.00 <sup>3</sup> |
| Maternal height, (cm) <sup>1</sup> |  | 160.7 (130-195) | 160.7 (130-195) | 160.4 (145-178) | 0.90 <sup>3</sup> |
| Body mass index, (kg/m<sup>2</sup>) <sup>1</sup> |  | 22.6 (11-71.1) | 22.6 (11-71.1) | 22.9 (15.4-50.3) | 0.93 <sup>3</sup> |
| Pregnancy times <sup>1</sup> |  | 2.4 (0-11) | 2.4 (0-11) | 2.7 (1-10) | 0.60 <sup>3</sup> |
| Delivery times <sup>1</sup> |  | 0.6 (0-4) | 0.6 (0-5) | 0.7 (0-4) | 0.84 <sup>3</sup> |
| **Adverse pregnancy outcomes in history and prenatal screening in current pregnancy** |  |  |  |  |  |
| **The adverse pregnancy outcomes of history** | Development stopped on embryonic <sup>2</sup> | 748 (1.15 %) | 730 (1.15 %) | 18 (1.08 %) | 0.89 <sup>4</sup> |
|  | Development stopped on fetal <sup>2</sup> | 4,048 (6.20 %) | 3,926 (6.17 %) | 122 (7.32 %) | 0.06 <sup>4</sup> |
|  | Spontaneous abortion <sup>2</sup> | 5,366 (8.22 %) | 5,201 (8.18 %) | 165 (9.90 %) | 0.01 <sup>4</sup> |
|  | Abortion <sup>2</sup> | 619 (0.95 %) | 597 (0.94 %) | 22 (1.32 %) | 0.14 <sup>4</sup> |
|  | **Summary for miscarriage <sup>2</sup>** | **10,319 (15.81%)** | **10,004 (15.73% )** | **315 (18.91%)** | **0.0006 <sup>4</sup>** |
|  | Molar pregnancy <sup>2</sup> | 130 (0.20 %) | 127 (0.20 %) | 3 (0.18 %) | 1.00 <sup>5</sup> |
|  | Biochemical pregnancy <sup>2</sup> | 580 (0.90 %) | 558 (0.90 %) | 22 (1.32 %) | 0.08 <sup>4</sup> |
|  | Extrauterine pregnancy <sup>2</sup> | 1,163 (1.80 %) | 1,117 (1.76 %) | 46 (2.76 %) | 0.003 <sup>4</sup> |
|  | **Summary for abnormal pregnancy <sup>2</sup>** | **1,841 (2.82%)** | **1,772 (2.79%)** | **69 (4.14%)** | **0.0012 <sup>4</sup>** |
|  | Others adverse pregnancy outcome | 646 (0.99 %) | 624 (0.98 %) | 22 (1.32 %) | 0.21 <sup>4</sup> |
|  | **Summary for adverse pregnancy outcomes <sup>2</sup>** | **12,404 (19.01 %)** | **12,015 (18.89 %)** | **389 (23.35 %)** | **5.53E-6 <sup>4</sup>** |
| **Prenatal screening in current pregnancy** | High risk of Down's syndrome in traditional screening (HRDS-T) <sup>2</sup> | 14,321 (21.95 %) | 13,983 (21.99 %) | 338 (20.29 %) | 0.10 <sup>4</sup> |
|  | High risk of Edward's syndrome in traditional screening (HRES-T) <sup>2</sup> | 667 (1.02 %) | 651 (1.02 %) | 16 (0.96 %) | 0.89 <sup>4</sup> |
|  | High risk of Neural tube defects in traditional screening (HRNTD) <sup>2</sup> | 403 (0.62 %) | 397 (0.62 %) | 6 (0.36 %) | 0.23 <sup>4</sup> |
|  | **Summary for multiples serum markers <sup>2</sup>** | **15312 (23.46%)** | **14953 (23.51%)** | **359 (21.55%)** | **0.07 <sup>4</sup>** |
|  | Multiple malformations <sup>2</sup> | 165 (0.25 %) | 159 (0.25 %) | 6 (0.36 %) | 0.32 <sup>4</sup> |
|  | Choroid plexus cysts <sup>2</sup> | 368 (0.56 %) | 362 (0.57 %) | 6 (0.36 %) | 0.36 <sup>4</sup> |
|  | Increased nuchal translucency or nuchal fold (I-NTF) <sup>2</sup> | 253 (0.39 %) | 241 (0.38 %) | 12 (0.72 %) | 0.04 <sup>4</sup> |
|  | Single umbilical artery and abnormal development on kidney and intestine (SADKI) <sup>2</sup> | 389 (0.60 %) | 378 (0.59 %) | 11 (0.66 %) | 0.85 <sup>4</sup> |
|  | Echogenic intracardiac focus <sup>2</sup> | 647 (0.99 %) | 639 (0.10 %) | 8 (0.48 %) | 0.04 <sup>4</sup> |
|  | Other ultrasound abnormal <sup>2</sup> | 232 (0.36 %) | 230 (0.36 %) | 2 (0.12 %) | 0.15 <sup>5</sup> |
|  | **Summary for ultrasound <sup>2</sup>** | **2054 (3.15 %)** | **2009 (3.16%)** | **45 (2.70 %)** | **0.32 <sup>4</sup>** |
|  | High risk of Down's syndrome by NIPS (HRDS-N) <sup>2</sup> | 299 (0.46 %) | 284 (0.45 %) | 15 (0.90 %) | 0.01 <sup>4</sup> |
|  | High risk of Edward's syndrome by NIPS (HRES-N) <sup>2</sup> | 89 (0.14 %) | 87 (0.14 %) | 2 (0.12 %) | 1.00 <sup>5</sup> |
|  | High risk of Patau's syndrome by NIPS (HRPS-N) <sup>2</sup> | 50 (0.08 %) | 48 (0.08 %) | 2 (0.12 %) | 0.37 <sup>5</sup> |
|  | **Summary for NIPS <sup>2</sup>** | **436 (0.67 %)** | **417 (0.66 %)** | **19 (1.14 %)** | **0.02 <sup>4</sup>** |
|  | **Summary for prenatal screening in current pregnancy <sup>2</sup>** | **17,468 (26.77%)** | **17,052 (26.82%)** | **416 (24.97%)** | **0.098 <sup>4</sup>** |

<sup>1</sup> It means that the mean, minimum and max used to descript the characteristics of adverse pregnancy outcomes and prenatal screening result features.
<sup>2</sup> It means that the number and percentage used to descript the characteristics of adverse pregnancy outcomes and prenatal screening result features.
<sup>3</sup> Estimated the distribution different between HBV negative and positive by T test.
<sup>4</sup> Estimated the distribution different between HBV negative and positive by $\chi^2$ .
<sup>5</sup> Estimated the distribution different between HBV negative and positive by Fisher's exact test.

## Result

According to the guideline of ACMG, 65,257 pregnant women performed NIPS in our hospital from July 2015 to Nov 2019. Based on the situation of maternal HBV infection, participant was categorized into the group of HBV-P (infected) and HBV-N (un-infected) and 1,666 (2.5%) and 63,591 (97.5%) pregnant women contained, respectively. The baseline characteristic and distributional difference of population characters, adverse pregnancy outcomes and prenatal screening results in the current pregnancy between HBV-N and HBV-P were shown in Table1. We noted that the spontaneous abortion, extrauterine pregnancy, I-NTF, echogenic intracardiac focus and HRDS-N have significant difference distribution between HBV-P and HBV-N, and that their preparation were respectively lower in HBV-N than HBV-P: (8.18% vs 9.9%, p=0.01), (1.76% vs 2.76%, p=0.003), (0.38% vs 0.72%, p=0.04), (0.1% vs 0.48%, p=0.04) and (0.45% vs 0.9%, p=0.01). Meanwhile, the significant differences were also observed in the summaries of miscarries (15.73% vs 18.91%, p=0.0006), abnormal pregnancy (2.79% vs 4.14%, p=0.0012), adverse pregnancy outcomes (18.89% vs 23.35%, p=5.53E-6) and NIPS (1.14% vs 0.66%, p=0.02) (Table 1). The results indicated that maternal HBV infection may be significantly associated with some adverse pregnancy outcomes and prenatal screening results.

To estimate the robustness and sensitivity of our results, the three kinds of multivariate logic regression model were performed to obtain the adjusted RRs and 95% CI of each adverse pregnancy outcomes and prenatal screening results for women with HBV infected (Table 2). In the most of investigated adverse pregnancy outcomes and prenatal screening, their risk did not increase in the women with HBV infected than un-infected, except biochemical pregnancy(aRRs: 1.54, 95%CI: 0.97-1.75 in model C), extrauterine pregnancy(aRRs:1.36, 95%CI: 1-1.82 in model C), HRDS-T (aRRs:1.16, 95%CI: 1.02-1.32 in full model), I-NTF (aRRs: 2.3, 95%CI: 1.21-3.94 in full model), and HRDS-N (aRRs:1.9, 95%CI:1.08-3.09 in full mode). The results revealed the risk of maternal HBV infection in the different stage of pregnancy. The higher risk of biochemical pregnancy and extrauterine pregnancy reflect the barrier of women with HBV infected on gamete transfer and embryo implantation disorder in early stage of pregnancy. The higher risks of HRDS-T, I-NTF and HRDS-N were shown the development disorder of fetus in the second-trimeste of gestation. In the multivariate logistic regression models of full model, the risk of women with HBV infected is higher 16%, 130%, and 90% on HRDS-T, I-NTF, HRDS-N than uninfected, respectively, though their detection methods were different. Furthermore, the maternal age as a necessary covariate was used in all multivariate logistic regression to keep the robustness of our result and independently analyzed the association between it and miscarriage (Fig 1B). The additional risk of miscarriage is 96% and 66% higher in the women whose age are over 35 (aRRs: 1.96, 95%CI:1.84-2.09) and between 30 and 35 (aRRs: 1.84, 95%CI:1.58-1.75) than whose age lower 30, respectively (Fig 1B). Meanwhile, the woman with HBV infection had additional 37%, 18%, 21%, 62% and 23% on the high risk of the summarize of abnormal pregnancy (aRRs: 1.37, 95%CI: 1.07-1.75 in model C) and adverse pregnancy outcomes (aRRs: 1.18, 95%CI: 1.05-1.33 in model C), MSM (aRRs:1.16, 95%CI:1.02-1.32 in full model), NIPS (aRRs:1.64, 95%CI:0.99-2.53 in full model) and prenatal screening results (aRRs:1.18, 95%CI:1.04-1.33 in full model) than un-infected (Table 2 and Fig 1 A).

**Table 2:**
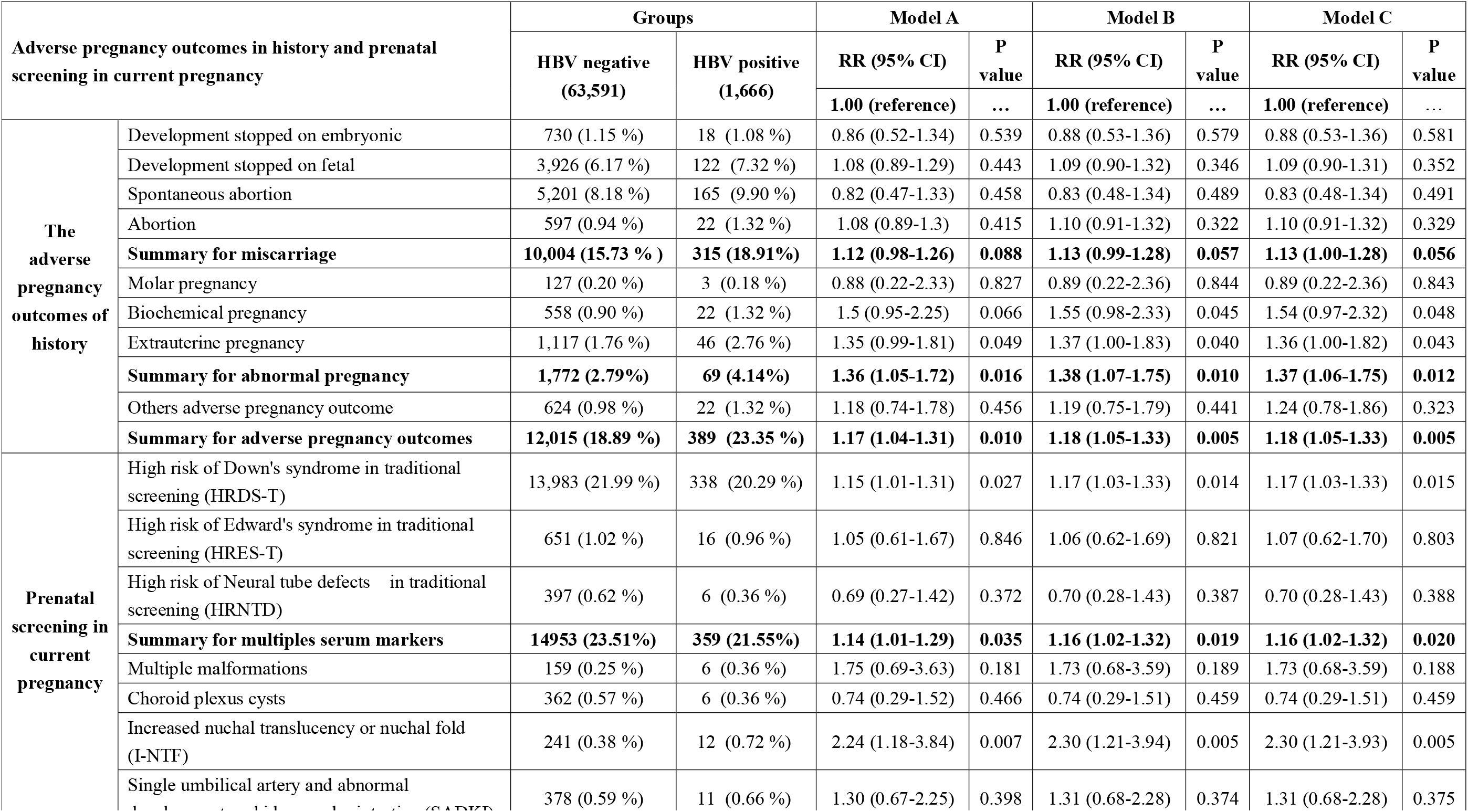
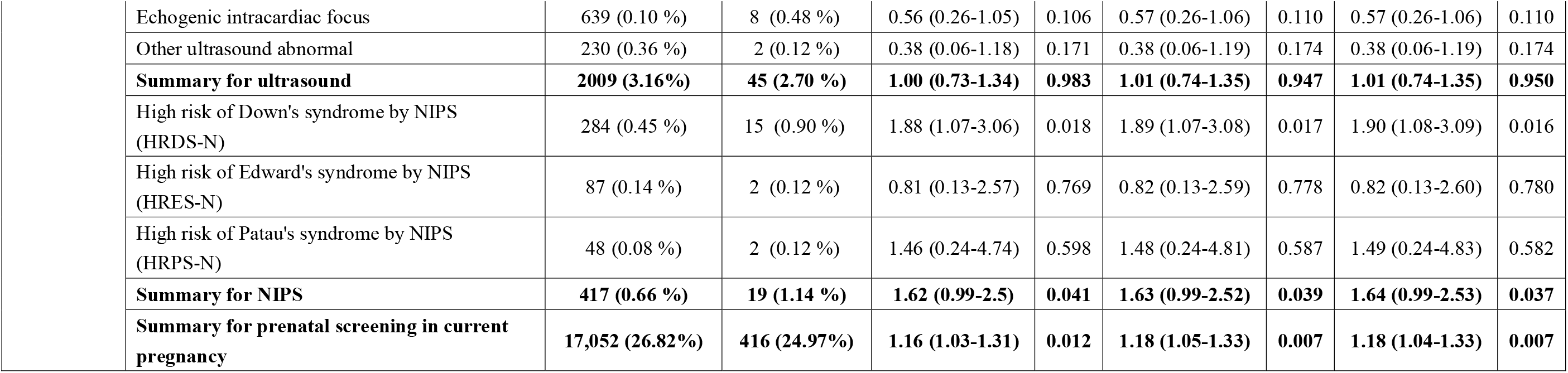
Association analysis between maternal HBV infection and risk of prenatal screening results in the multivariate model of A, B and C.

| Adverse pregnancy outcomes in history and prenatal screening in current pregnancy |  | Groups |  | Model A |  | Model B |  | Model C |  |
| --- | --- | --- | --- | --- | --- | --- | --- | --- | --- |
|  |  | HBV negative<br>(63,591) | HBV positive<br>(1,666) | RR (95% CI) | P<br>value | RR (95% CI) | P<br>value | RR (95% CI) | P<br>value |
|  |  |  |  | 1.00 (reference) | ... | 1.00 (reference) | ... | 1.00 (reference) | ... |
| The adverse pregnancy outcomes of history | Development stopped on embryonic | 730 (1.15 %) | 18 (1.08 %) | 0.86 (0.52-1.34) | 0.539 | 0.88 (0.53-1.36) | 0.579 | 0.88 (0.53-1.36) | 0.581 |
|  | Development stopped on fetal | 3,926 (6.17 %) | 122 (7.32 %) | 1.08 (0.89-1.29) | 0.443 | 1.09 (0.90-1.32) | 0.346 | 1.09 (0.90-1.31) | 0.352 |
|  | Spontaneous abortion | 5,201 (8.18 %) | 165 (9.90 %) | 0.82 (0.47-1.33) | 0.458 | 0.83 (0.48-1.34) | 0.489 | 0.83 (0.48-1.34) | 0.491 |
|  | Abortion | 597 (0.94 %) | 22 (1.32 %) | 1.08 (0.89-1.3) | 0.415 | 1.10 (0.91-1.32) | 0.322 | 1.10 (0.91-1.32) | 0.329 |
|  | **Summary for miscarriage** | **10,004 (15.73 % )** | **315 (18.91%)** | **1.12 (0.98-1.26)** | **0.088** | **1.13 (0.99-1.28)** | **0.057** | **1.13 (1.00-1.28)** | **0.056** |
|  | Molar pregnancy | 127 (0.20 %) | 3 (0.18 %) | 0.88 (0.22-2.33) | 0.827 | 0.89 (0.22-2.36) | 0.844 | 0.89 (0.22-2.36) | 0.843 |
|  | Biochemical pregnancy | 558 (0.90 %) | 22 (1.32 %) | 1.5 (0.95-2.25) | 0.066 | 1.55 (0.98-2.33) | 0.045 | 1.54 (0.97-2.32) | 0.048 |
|  | Extrauterine pregnancy | 1,117 (1.76 %) | 46 (2.76 %) | 1.35 (0.99-1.81) | 0.049 | 1.37 (1.00-1.83) | 0.040 | 1.36 (1.00-1.82) | 0.043 |
|  | **Summary for abnormal pregnancy** | **1,772 (2.79%)** | **69 (4.14%)** | **1.36 (1.05-1.72)** | **0.016** | **1.38 (1.07-1.75)** | **0.010** | **1.37 (1.06-1.75)** | **0.012** |
|  | Others adverse pregnancy outcome | 624 (0.98 %) | 22 (1.32 %) | 1.18 (0.74-1.78) | 0.456 | 1.19 (0.75-1.79) | 0.441 | 1.24 (0.78-1.86) | 0.323 |
|  | **Summary for adverse pregnancy outcomes** | **12,015 (18.89 %)** | **389 (23.35 %)** | **1.17 (1.04-1.31)** | **0.010** | **1.18 (1.05-1.33)** | **0.005** | **1.18 (1.05-1.33)** | **0.005** |
| Prenatal screening in current pregnancy | High risk of Down's syndrome in traditional screening (HRDS-T) | 13,983 (21.99 %) | 338 (20.29 %) | 1.15 (1.01-1.31) | 0.027 | 1.17 (1.03-1.33) | 0.014 | 1.17 (1.03-1.33) | 0.015 |
|  | High risk of Edward's syndrome in traditional screening (HRES-T) | 651 (1.02 %) | 16 (0.96 %) | 1.05 (0.61-1.67) | 0.846 | 1.06 (0.62-1.69) | 0.821 | 1.07 (0.62-1.70) | 0.803 |
|  | High risk of Neural tube defects in traditional screening (HRNTD) | 397 (0.62 %) | 6 (0.36 %) | 0.69 (0.27-1.42) | 0.372 | 0.70 (0.28-1.43) | 0.387 | 0.70 (0.28-1.43) | 0.388 |
|  | **Summary for multiples serum markers** | **14953 (23.51%)** | **359 (21.55%)** | **1.14 (1.01-1.29)** | **0.035** | **1.16 (1.02-1.32)** | **0.019** | **1.16 (1.02-1.32)** | **0.020** |
|  | Multiple malformations | 159 (0.25 %) | 6 (0.36 %) | 1.75 (0.69-3.63) | 0.181 | 1.73 (0.68-3.59) | 0.189 | 1.73 (0.68-3.59) | 0.188 |
|  | Choroid plexus cysts | 362 (0.57 %) | 6 (0.36 %) | 0.74 (0.29-1.52) | 0.466 | 0.74 (0.29-1.51) | 0.459 | 0.74 (0.29-1.51) | 0.459 |
|  | Increased nuchal translucency or nuchal fold (I-NTF) | 241 (0.38 %) | 12 (0.72 %) | 2.24 (1.18-3.84) | 0.007 | 2.30 (1.21-3.94) | 0.005 | 2.30 (1.21-3.93) | 0.005 |
|  | Single umbilical artery and abnormal development on kidney and intestine (SADKI) | 378 (0.59 %) | 11 (0.66 %) | 1.30 (0.67-2.25) | 0.398 | 1.31 (0.68-2.28) | 0.374 | 1.31 (0.68-2.28) | 0.375 |

|  |  |  |  |  |  |  |  |  |
| --- | --- | --- | --- | --- | --- | --- | --- | --- |
| Echogenic intracardiac focus | 639 (0.10 %) | 8 (0.48 %) | 0.56 (0.26-1.05) | 0.106 | 0.57 (0.26-1.06) | 0.110 | 0.57 (0.26-1.06) | 0.110 |
| Other ultrasound abnormal | 230 (0.36 %) | 2 (0.12 %) | 0.38 (0.06-1.18) | 0.171 | 0.38 (0.06-1.19) | 0.174 | 0.38 (0.06-1.19) | 0.174 |
| **Summary for ultrasound** | **2009 (3.16%)** | **45 (2.70 %)** | **1.00 (0.73-1.34)** | **0.983** | **1.01 (0.74-1.35)** | **0.947** | **1.01 (0.74-1.35)** | **0.950** |
| High risk of Down's syndrome by NIPS (HRDS-N) | 284 (0.45 %) | 15 (0.90 %) | 1.88 (1.07-3.06) | 0.018 | 1.89 (1.07-3.08) | 0.017 | 1.90 (1.08-3.09) | 0.016 |
| High risk of Edward's syndrome by NIPS (HRES-N) | 87 (0.14 %) | 2 (0.12 %) | 0.81 (0.13-2.57) | 0.769 | 0.82 (0.13-2.59) | 0.778 | 0.82 (0.13-2.60) | 0.780 |
| High risk of Patau's syndrome by NIPS (HRPS-N) | 48 (0.08 %) | 2 (0.12 %) | 1.46 (0.24-4.74) | 0.598 | 1.48 (0.24-4.81) | 0.587 | 1.49 (0.24-4.83) | 0.582 |
| **Summary for NIPS** | **417 (0.66 %)** | **19 (1.14 %)** | **1.62 (0.99-2.5)** | **0.041** | **1.63 (0.99-2.52)** | **0.039** | **1.64 (0.99-2.53)** | **0.037** |
| **Summary for prenatal screening in current pregnancy** | **17,052 (26.82%)** | **416 (24.97%)** | **1.16 (1.03-1.31)** | **0.012** | **1.18 (1.05-1.33)** | **0.007** | **1.18 (1.04-1.33)** | **0.007** |

**Figure 1:**
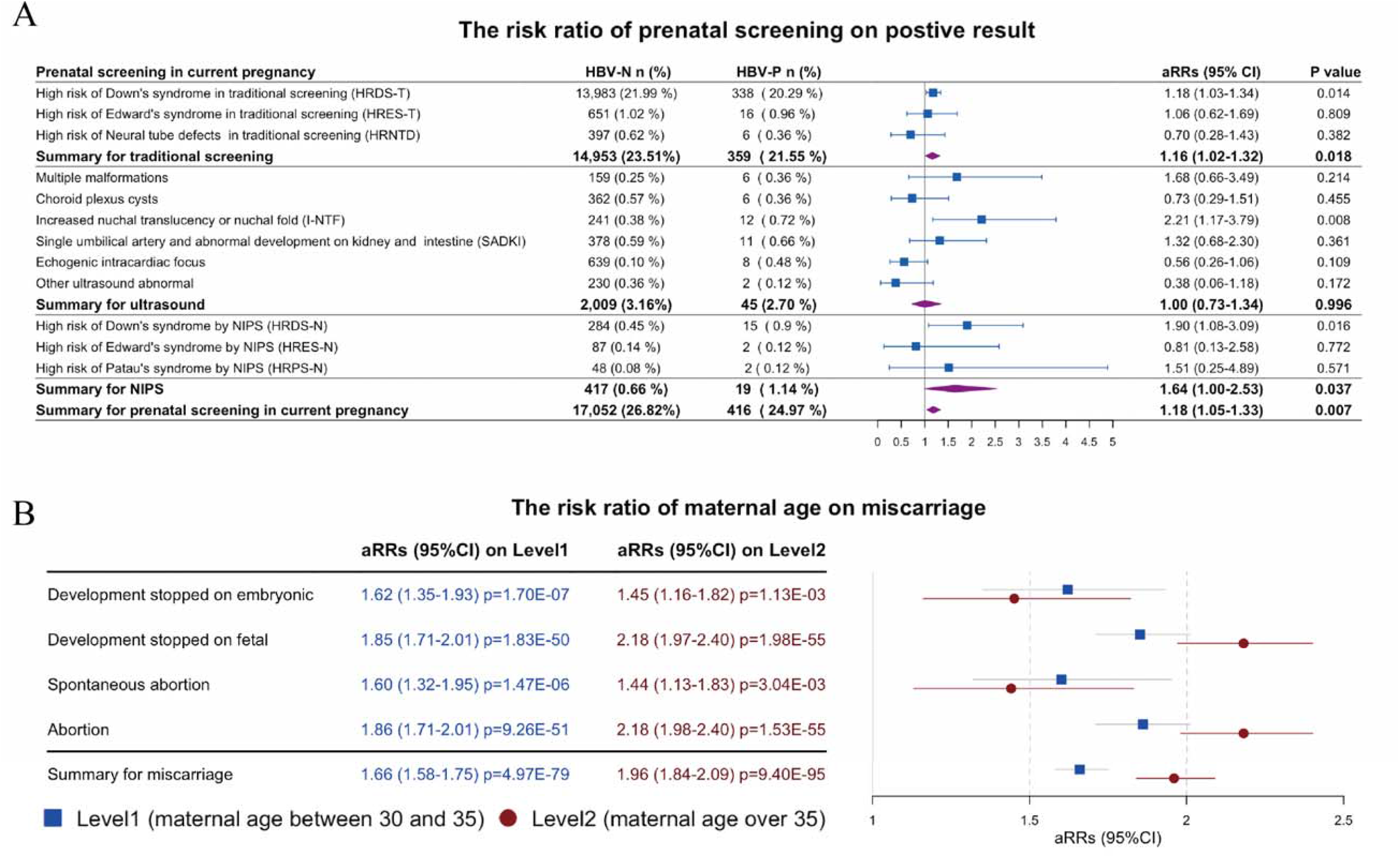
The risk ratio of women with HBV infection. A) The risk ratio of maternal HBV infection on prenatal screening results using logistic multivariate regression in full model. B) The risk ratio on the different stage of maternal age in miscarriages using logistic multivariate regression in model C.

Furthermore, 299 pregnancy women who reported DS high risk in NIPS were confirmed by amniocentesis or follow-up in which 284 were from HBV-N and 15 were from HBV-P (Fig 2A). According to their diagnostic results or follow-up information, they were categorized into five classes: conformed by amniocentesis, abortion without diagnosis, fetal development stopped, refusing diagnoses and false-positive results (Fig 2A). The true positive ratio (TPR) of DS in HBV-P (86.7% (13/15)) is over HBV-N (80.6% (229/284)) and the incidence of DS in HBV-P (0.78% (13/1,666)) is twice than HBV-N (0.36% (229/63,519)). The difference of true positive results between HBV-P and HBV-N examined by Fisher’s exact test and significant (p=0.012) (Fig 2B). Meanwhile, The aRRs and 95% CI in model A(aRRs:2.01,95%CI:1.09-3.39), B (aRRs:1.99, 95%CI:1.08-3.34), C (aRRs:1.99, 95%CI:1.08-3.34) and full (aRRs:1.98, 95%CI: 1.07-3.34) were significant and supported that maternal HBV infection is an independent risk factor of fetal Down syndrome (Fig 2C). To assess comprehensively the impact of maternal HBV infection on fetal DS, we proposed that the screening result of high risk on DS in NIPS-N group were truly, and calculated the difference between women with HBV infected and un-infected after accumulated them step by step (Fig 2B and C). The result also suggested that women with HBV infection have significant additional risk than women with HBV un-infected. Additionally, the different of maternal age in distribution was also estimated as an independent factor by T-test but their different were not significant (Fig 2D).

**Figure 2:**
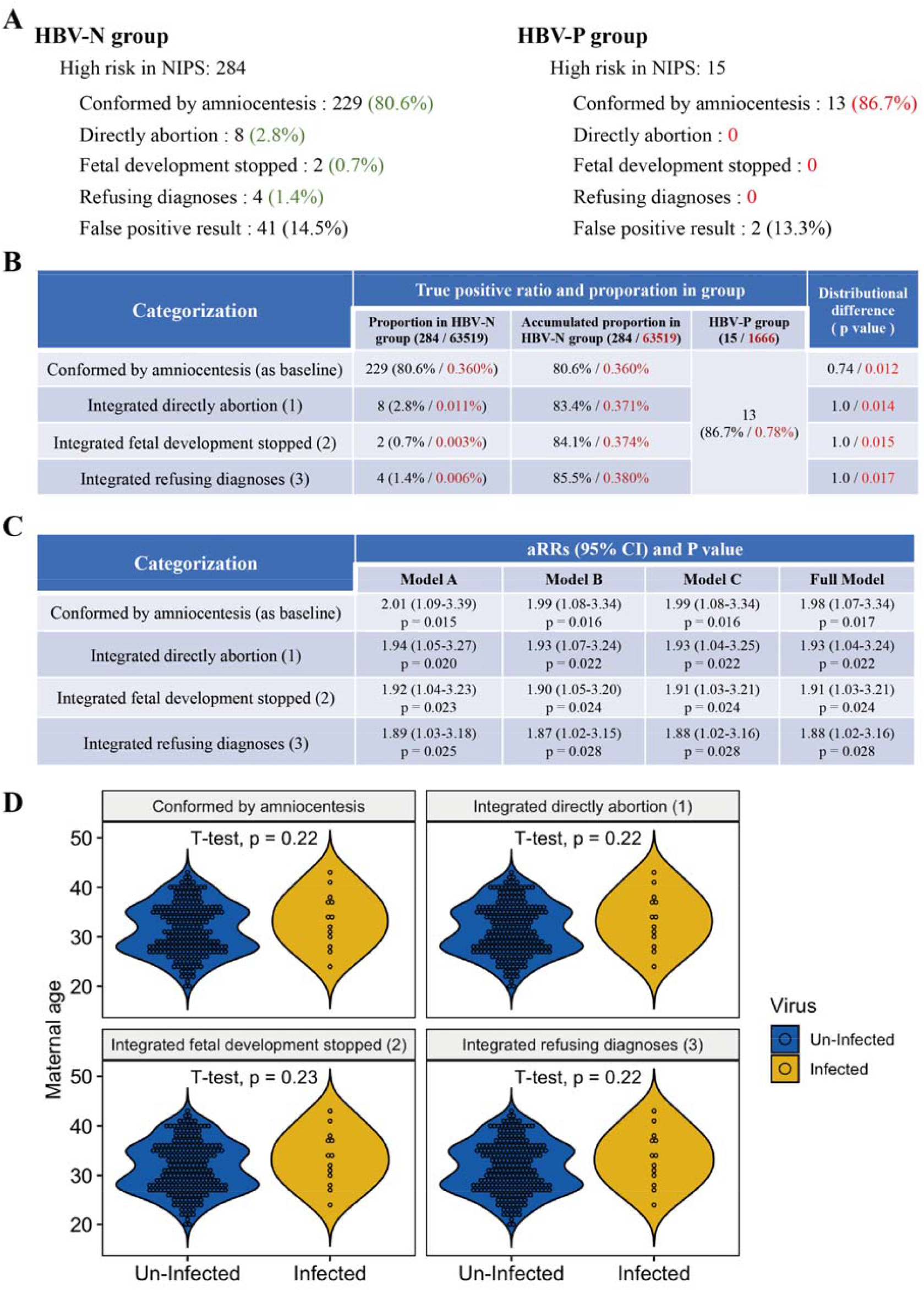
Diagnostic result for women with the positive result of fetal Down’s syndrome in NIPS. A) The categorization of diagnosing for women with high risk of fetal Down’s syndrome in HBV-P and HBV-N. Their proportion were marked by red and green, respectively. In B and C, we proposed that the cases of directly abortion, fetal development stopped and refusing diagnose were conform by amniocentesis as truly positive results. B) The proportion of diagnosed and accumulated for the cases with high risk of fetal DS in NIPS were marked by black; the red marked them in the group of HBV-N and HBV-P. The p value demonstrated the distributional difference of accumulated diagnose ratio in HBV-N and HBV-P by χ2. The accumulated order marked by number and followed in the name of categorization. C) The risk ratio and 95% CI of fetal DS in women with HBV infected estimated after accumulated the diagnose number. D) The distributional difference of maternal age with HBV infected and un-infected between HBV-N and HBV-P group and the number in subfigure titles marked the integrated order.

## Discussion

According to the collection information from 69,527 pregnant women, we investigated the association between maternal HBV infection and eight kinds of adverse pregnancy outcomes and 12 kinds of prenatal screening results. Our results suggested that the maternal HBV infection was significantly associated with the biochemical pregnancy, extrauterine pregnancy and fetal DS. The high risk of fetal DS shown at the three results of HRDS-T, HRDS-N and I-NTF in the prenatal screening of MSM, ultrasound and NIPS, respectively. The fetus of women with HBV infected were more likely to be reported as positive result of DS in MSM, ultrasound and NIPS than the fetus of women with HBV un-infected. Meanwhile, the diagnostic result of amniocentesis from the positive result of DS in NIPS also conformed the additional risk of fetal DS in the women with HBV infection and twice than the fetus from the women with HBV un-infected. Furthermore, the result also indicated that the influence of maternal HBV infection across different stage because the additional risk of biochemical pregnancy and extrauterine pregnancy reflect the barrier of women with HBV infected on gamete transfer and embryo implantation disorder in early stage of pregnancy; the higher risks of HRDS-T, I-NTF and HRDS-N shown the development disorder of fetus in the second-trimeste of gestation. Our study provides direct and strong evidence to reveal the risk of maternal HBV infection in gestation and indicate that HBV infection may not only influence the maternal protein expression (such as AFP) but also disturb the genomic stabilization of gamete and fetus. However, we also noted that Edward’s syndrome and Patau’s syndrome were not associated with maternal HBV infection which may be due to the smaller number of positive cases or chromosomes different on sequence of chromosome 18 and 13 with chromosome 21.

Miscarriage is a common adverse pregnancy outcome, and the cause of it has been widely discussed in previous researches. According to the health information from Shoklo Malaria Research Unit (SMRU), Marieke Bierhoff and colleagues suggested that maternal HBV infection is not significantly associated with miscarriage after investigating 11,025 women from the Myanmar-Thailand border (Southeast Asia)^33^. The odds ratio (OR) of miscarriage between HBsAg-/HBeAg- and HBsAg+/HBeAg-is not significant (OR:1.23, 95%CI is 0.88-1.71, p=0.226), as well as between HBsAg-/HBeAg- and HBsAg+/HBeAg+ (OR:0.94, 95%CI: 0.71-1.25, p= 0.690). Linlin Wang *et al* reported the same result with Marieke Bierhoff *et al* after investigation 8,550 infertile patients with treatment by vitro fertilization (miscarriage ratio in HBsAg^+^/HBeAg^+^ [11.7%], HBsAg^+^/HBeAg^-^ [10.0%], HBsAg^-/^HBeAg^-^ [11.7%])^34^. However, we also noted that the miscarriage risk increased by 50%^17^ in the pregnant women with more advanced diseases due to HBV infection, such as cirrhotic. In clinical, the doctors always emphasize women’s health on the stage of pre-pregnancy and pregnancy for protecting women, fetuses health and decreasing the potential risk of adverse pregnancy outcomes. Thus, the pregnant women with advanced diseases for HBV infection were almost not seen in our cohort though the concentration of HBV DNA or HBV envelope antigen in serum were not tested. Gang Qin *et al*. reported maternal HBV infection is a risk factor of miscarriage with the pregnant women of advanced age after investigation 21,004 pregnant women^12^. Maternal age is an inherent sensitivity factor of miscarriage^35,36^, even without HBV infection, the higher rate of miscarriage was also observed in the pregnancy women with advanced age than young ^37,38^. The sample size of retrospective cohort in our research is over three times than Gang Qin *et al’s* study, which improved the validation and reliability of statistics.

In the twelve prenatal screening results, three of them were significantly associated with fetal Down’s syndrome with maternal HBV infection. MSM is a traditional method to assess fetal Down’s syndrome risk in second-trimester by protein quantification. However, the sensitivity and specificity of MSM depended on target protein, sampling environment and local cut off value, as well as the association between maternal HBV infection and the screening result is rare report. In Po-Jen Cheng *et al* research, they suggested that the maternal HBV infection is not associated with the screening result of MSM after investigated 8,193 eligible pregnancies^39^. In their study, the target protein of marking fetal DS is pregnancy-associated plasma protein-A (PAPP-A), not protein AFP. Many years ago, protein AFP was considered as a valuable diagnostic and prognosis prediction biomarker in hepatocellular cancer due to HBV infection^40,41^. Thus, these two results are not be used comparing (Po-Jen Cheng *et al* research and our study) and indicated that protein AFP is sensitive with HBV infection than protein PAPP-A.

In our study, the main limitation is that the HBV infection status of women was not estimated. The DNA and envelope antigen concentration of HBV were important risk factors of vertical transmission^42-44^ and key indicators of fetal immunization with HBV. The information about HBV envelope antigen or HBV DNA also not contain in the application documents of NIPS and followed up. The neonatal information is missing, such as gestation week, fetal weight and height and the complication of maternal during delivery. They were made it hard to further assess the influence of maternal HBV infection with viral transmission, neonatal outcomes and administration of preventive antiviral therapy.

In summary, our study suggested that women infected with HBV have significant additional risk in biochemical pregnancy, extrauterine pregnancy and the fetal Down’s syndrome. It provides new evidence to reveal the pregnant risk of women with HBV infected and suggested the detail category of clinical phonotype will be helpful to understand the impact of pregnancy with maternal HBV infection. Thus, we suggested that HBV infection detection and appropriate medical intervention in pre-pregnancy were necessary and helpful to prevent fetal Down’s syndrome, other unknow pregnant risk and adverse pregnancy outcomes.

## Data Availability

The datasets generated during and/or analysed during the current study are available from the corresponding author on reasonable request.

## Contributors

Quanze He searched the literature, designed the study, analyzed the data, interpreted the results, and drafted the manuscript. You Zhou improved the manuscript and interpreted the results together with Hankui Liu, Xiao Dang, and Jianguo Zhang. Xiaojuan Wu, Ying Xue, Chunhua Zhang and Lu Lu collected the data and revised the manuscript. Tim Wang and Hong Li conceived, designed, and supervised the study, interpreted the results, and revised the manuscript. All authors contributed to the writing of the manuscript.

## Declaration of interests

We declare no competing interests.

## Acknowledgments

This work is supported by Jiangsu Provincial Commission of Health and Family Planning Research Project (X201604), Jiangsu Maternal and Children health care key discipline (FXK201748), Jiangsu Maternal and Children health care research project (F201603), Jiangsu Provincial Medical Innovation Team (CXTDB2017013), Suzhou Key Medical Center (SZZX201505), Suzhou Introduced Project of Clinical Medical Expert Team (SZYJTD201708) regression in full model. B) The risk ratio on the different stage of maternal age in miscarriages using logistic multivariate regression in model C.

## Summary

### What is already known about this subject?

1. Maternal HBV infection has been considered to be associated with adverse pregnancy outcomes but the result from some studies were controversial, even in the cohort studies with large sample size.
2. Down syndrome is a rare disease and the higher infection rate of HBV was observed in the patient of DS than other patient of nervous system diseases and the normals.
3. Contact with blood or other body fluids in childbirth is an important transmission channel of HBV from mother to child.

### What are the new findings?

1. Maternal HBV infection was an independent risk factor of fetal Down syndrome, biochemical pregnancy and extrauterine pregnancy.
2. Maternal HBV infection is a potential regulator of maternal protein expression and may affect the stabilization of fetal chromosome.

### How might it impact on clinical practice in the foreseeable future?

1. HBV infection detection and appropriate medical intervention in pre-pregnancy will be necessary and helpful to prevent fetal Down’s syndrome, other unknown pregnant risk and adverse pregnancy outcomes.
2. There is a need to investigate the pathogenesis of biochemical pregnancy, extrauterine pregnancy and fetal Down syndrome by in the future.

## Notes

### Competing Interest Statement

The authors have declared no competing interest.

